# Reduced COVID-19-Related Critical Illness and Death, and High Risk of Epidemic Resurgence, After Physical Distancing in Ontario, Canada

**DOI:** 10.1101/2020.04.29.20084475

**Authors:** Ashleigh R. Tuite, Amy L. Greer, Steven De Keninck, David N. Fisman

## Abstract

**Background:** Insights from epidemiological models have helped to both guide and better understand COVID-19 mitigation policies that have been adopted across the globe. Many early models focussed on initial control options and were less reliant on fitting to observed data. As the pandemic progresses, models can be used to quantify the impact that control measures have had and what may unfold when such measures are relaxed.

**Objective:** To explore the impact of physical distancing measures on COVID-19 transmission in the population of Ontario, Canada.

## Methods and Findings

We used a previously described age- and health-status stratified transmission model of COVID-19 in the Canadian province of Ontario (1). Parameters for the latent period and pre-symptomatic period were updated based on more recent data (2). We assumed that 10% of mild/moderate infections were detected and isolated, and 10% of exposed cases were quarantined. We also assumed a 70% reduction in contacts with the implementation of physical distancing measures, which came into effect approximately 3 weeks after the model start date of 6 March 2020.

The model was fit using maximum likelihood estimation. We allowed the following parameters to vary during fitting: basic reproductive number (R_0_), initial number of infected individuals; infectious period, and average length of stay in ICU. All other parameters were unchanged from (1).

We fit the model to confirmed cases occupying intensive care unit (ICU) beds and COVID-19 attributable deaths among hospitalized cases for the time period 19 March - 26 April 2020 **(Figure 1)**. We compared the fitted model to a eounterfaetual scenario where physical distancing was not introduced. Without intervention, we projected up to 33.4 (95% credible interval (CrI): 14.3 – 73.8) cases in ICU per 100,000 population, compared to 2.0 (95% CrI: 1.4–3.0) cases per 100,000 with physical distancing. Similarly, the number of deaths among hospitalized cases without intervention (10.3 (95% CrI: 5.7–19.9 per 100,000) was projected to be 4.6-fold what was observed with physical distancing (2.2 (95% CrI: 1.7–3.0) per 100,000).

**Figure 1.**
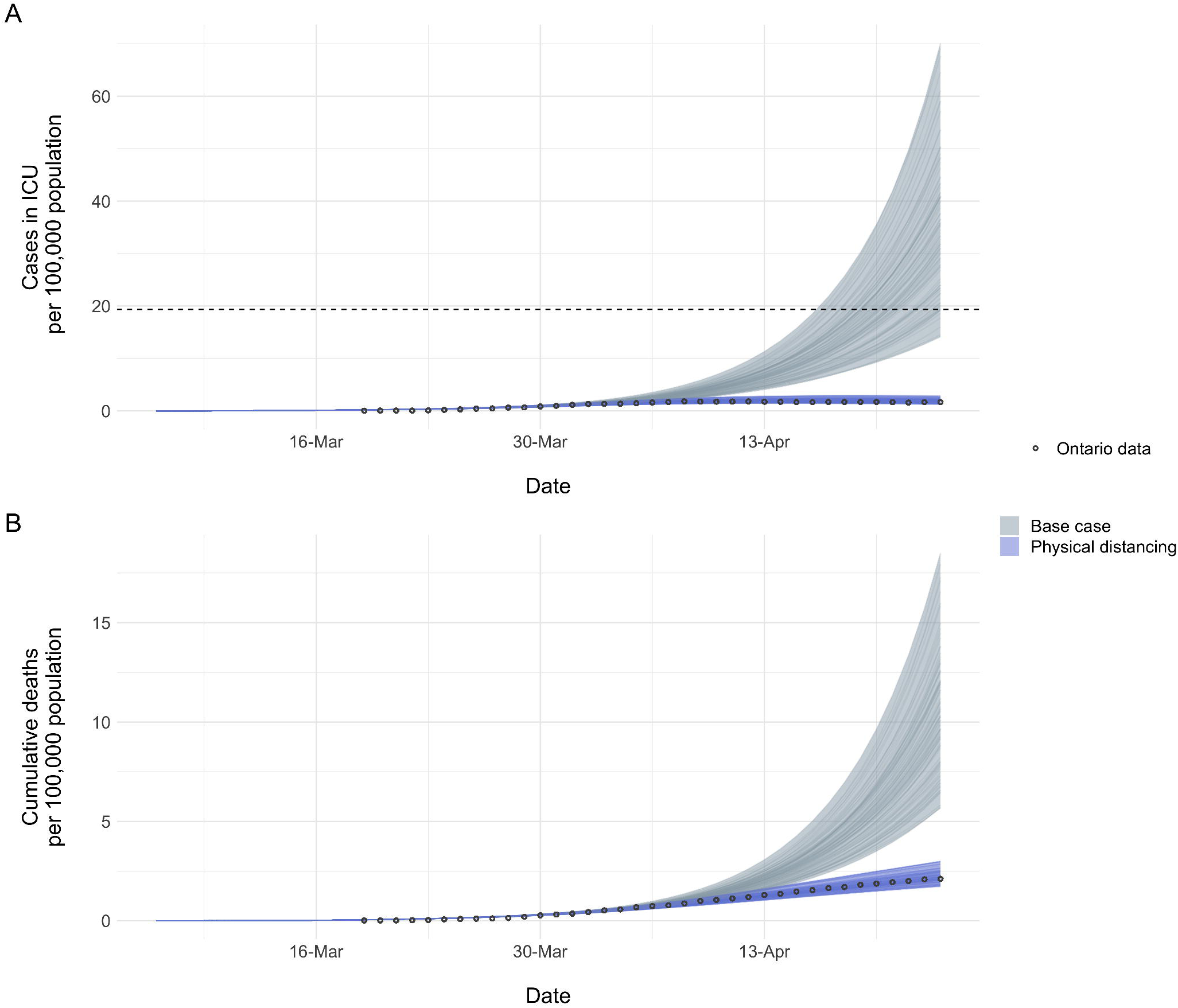
Model-projected COVID-19 outcomes with and without physical distancing measures. (A) Prevalent cases in ICU and (B) cumulative deaths are shown in the presence of physical distancing, which is assumed to reduce contacts to 30% of normal. Deaths exclude those occurring outside of the hospital (e.g., in long term care facilities). Bands represent the 95% credible intervals derived from 100 model simulations per scenario. The horizonal dashed lined in (A) represents total ventilated ICU beds (19.3) per 100,000 population in Ontario as a measure of maximum ICU capacity. Fitted parameter values were as follows: R_o_ (2.97); initial number of infected individuals (670); infectious period (4 days); average length of stay in ICU (15 days).

Relaxation of physical distancing measures without compensatory increases in case detection, isolation, and/or contact tracing was projected to result in a resurgence of disease activity. The return to normal or near-normal levels of contact would rapidly result in cases exceeding ICU capacity **(Figure 2)**. Maintaining physical distancing for a longer period of time, allowing for the initial wave of infections to subside, delayed this resurgence, but the level of contacts post-restrictive distancing was the major factor determining how quickly ICU capacity was expected to be overwhelmed.

**Figure 2.**
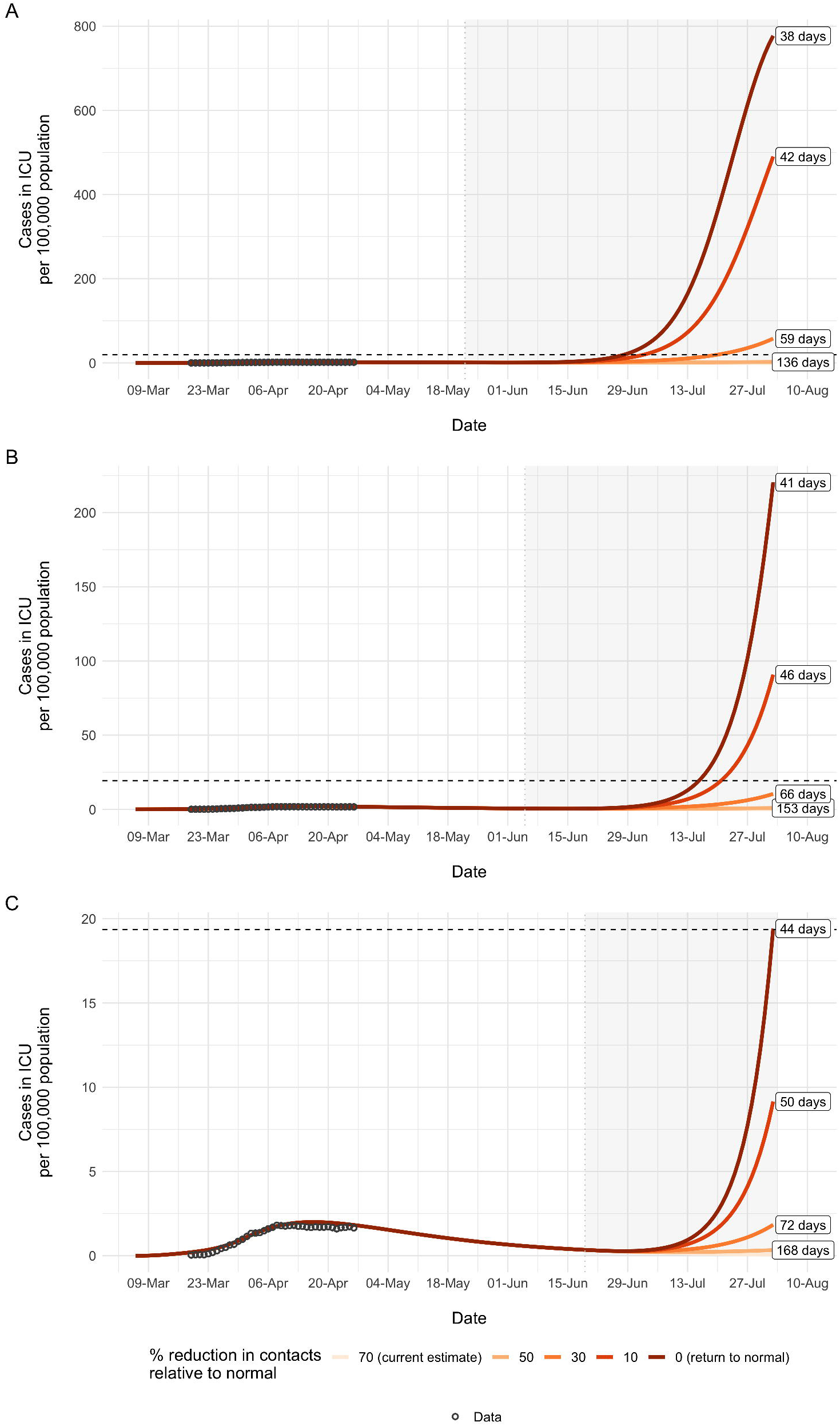
Effect of relaxation of physical distancing measures on projected ICU requirements. Results are shown for (A) 8-week, (B) 10-week, and (C) 12-week periods of physical distancing prior to relaxation of distancing measures. The grey shaded area represents the time period during which contact rates were allowed to increase. Prior to this, restrictive physical distancing measures were in place, with contacts reduced by 70% of normal. Baseline contact rates (without physical distancing) were derived from (5). Maximum ICU bed capacity in Ontario is indicated by the dashed horizontal line (19.3 vented beds per 100,000 population). The time until ICU capacity is exceeded after relaxation of physical distancing measures is indicated by the labels. Note that for some scenarios, the time until ICU capacity is surpassed extended beyond the time scale shown in the graphs; we restricted the x-axis to aid with interpretability among the scenarios. If contacts remain at 70% of normal ICU capacity is not projected to be exceeded, so no labels appear for this scenario.

## Discussion

To date in Ontario, the number of cases in ICU has remained below current (recently expanded) ICU capacity. The provincial response was initiated in mid-March, with the declaration of a state of emergency on March 17,2020. Without intervention, we projected that Ontario would have rapidly exceeded ICU capacity and observed substantially higher mortality. Our modeling also demonstrates the challenges associated with relaxation of physical distancing measures without a concomitant increase in other public health measures. Specifically, when the number of contacts between individuals return to more than 50% of normal, we expect rapid resurgence of disease activity and ICUs quickly reach capacity.

A limitation of our model is that it was fit to mortality among hospitalized cases and the results presented here apply to community transmission. Ontario, as in many other jurisdictions, is experiencing outbreaks in long-term care homes (LTCH). 65% of confirmed COVID-19 deaths in the province to date have occurred outside of hospitals and there is a divergence between trends in hospitalizations and mortality that represents different pathways of care for individuals in LTCH (3, 4). Understanding and describing the dynamics of SARS-CoV-2 transmission in LTCH and other institutional settings is important for protecting the most vulnerable in our society and requires alternate modeling approaches and control measures.

We show deterministic outputs for the epidemic projections with different levels of relaxation of physical distancing. Given variability in R_o_, it is possible that local community transmission may be eliminated or time to resurgence will delayed. However, as long as SARS-CoV-2 is circulating globally, population susceptibility remains, and we have open borders, the risk of re-introduction and resurgence remains. Our results show the challenges that lie ahead as we move to the de-escalation phase of the first wave of the pandemic.

## Data Availability

All data are publicly available.

